# Healthcare utilisation survey as part of the Hybrid model of the Surveillance of Enteric Fever in India (SEFI) study: processes, results, and challenges

**DOI:** 10.1101/2021.02.27.21252424

**Authors:** Reshma Raju, J Kezia Angelin, Arun S Karthikeyan, Dilesh Kumar, Ranjith R Kumar, Nikhil Sahai, Karthik Ramanujan, Manoj Murhekar, A. Elangovan, Prasanna Samuel, Jacob John, Gagandeep Kang

## Abstract

**Background:** Enteric fever is a significant health challenge in low and middle-income countries. “Surveillance of Enteric Fever in India” (SEFI) network was set up to obtain reliable incidence data. Six sites utilised a hybrid surveillance model, a combination of facility-based surveillance and community-based healthcare utilisation survey (HCUS). HCUS was performed to determine the percentage of the catchment population utilising the study facilities for febrile episodes, and is described here.

**Methods:** A two-stage sampling process was utilised for the HCUS to select 5000 households per site. Demographic data and healthcare-seeking behaviour were assessed.

**Results:** Febrile hospitalisation rate ranged from 2.5/1000 in Kullu to 9.6/1000 in Anantapur. The percentage of febrile admissions that sought care in the study hospital from the catchment area is 17% in Anantapur, 38% in Karimganj, 38% in Chandigarh, 10% in Nandurbar, 36% in Kullu and 24% in East Champaran respectively.

**Conclusions:** The variability in healthcare utilisation for fever admissions in the study hospitals underscores the importance of periodic healthcare utilisation surveys in hybrid surveillance. It is essential to adjust for those cases which have slipped out of the facility surveillance radar for obtaining an accurate estimate of the disease burden.

## Background

The need for effective surveillance systems for assessing the burden of any disease has long been recognised. The WHO propounds that ‘effective communicable disease control relies on effective response systems and effective response systems rely on effective disease surveillance” [1–3]. Emerging events of public concern of a population vary from site to site and from time to time. Diseases that are common in one part of the globe at a point in time may be conspicuously absent in another. A rigorous community-based health monitoring and surveillance system are essential in the early detection of diseases, to facilitate informed policy decisions [4].

However reliable community-based data is scarce in low- and middle-income countries (LMICs) [5,6], and, hence policymakers are forced to rely on information collected by Healthcare facilities [4]. Nevertheless, not everyone who is ill, especially the underprivileged and the vulnerable, seek treatment due to numerous socio-cultural and health literacy factors [7]. Of those who decide to seek treatment, a significant proportion may prefer to rely on private practitioners, traditional healers, or local pharmacies for treatment [8,9]. The existing weak Health Management Information System (HMIS) in the country further compounds this issue as it frequently fails to capture disease events [10]. Hence facility-based surveillance alone would underestimate the disease burden because they provide neither an actual nor a representative picture of the health challenges of the communities [3,11].

Community-based surveys may yield more representative burden estimates but have their limitations too. With the already existing constraints of trained personnel and financial resources in LMICs, genuine efforts to monitor and measure health indicators on a large scale at the community level, maybe further challenged by language, social and geographic barriers [4,12]. Even though community-based surveys, when implemented well, provide a high degree of sensitivity, it avowedly lacks the specificity of sophisticated laboratory tests to confirm the diagnoses [12].

A hybrid surveillance model may be utilised to bridge the gap between facility-based surveillance and community-based surveys. Such a model would use a sentinel healthcare facility-based surveillance to calculate a crude incidence rate. This rate would then be adjusted for those cases that utilised other facilities for healthcare, obtained via annual/biannual healthcare utilisation surveys taking place in the catchment populations of the study facilities [13–18].

To generate data on the incidence and burden of severe enteric fever in India, a condition that remains a significant public health concern in the country, the Surveillance for Enteric Fever in India (SEFI) network was set up in 18 sites in the year 2017. Six of these sites utilised a hybrid surveillance model (2018-2020) for burden estimation. This paper describes the conduct and results of the community-based healthcare utilisation surveys that occurred at each of these sites, as part of the SEFI study.

The main aim of the study was to estimate the proportion of febrile hospitalisations that sought inpatient care at the SEFI tier 2 study hospital sites, from among their catchment populations.

## Methods

### Study setting

The study was designed as part of the SEFI network, a more extensive network of surveillance initiated to obtain the burden of enteric fever in India.

SEFI was conceptualised as a multi-tiered surveillance system aimed at estimating the burden of enteric fever at different rural, urban, slum, hilly and tribal settings across India with varying designs of study at each of the tiers. The hybrid surveillance model, a relatively newly developed, cost-effective and sustainable approach, was followed by the second tier. This hybrid surveillance was conducted in secondary hospital settings of five rural and one urban site and their catchment areas across India, viz., Rural Development Trust Hospital Bathalapalli (Anantapur district of Andhra Pradesh), Makunda Christian Leprosy and General Hospital, Bazaricherra (Karimganj district of Assam), Duncan Hospital Raxaul (East Champarna district of Bihar), Lady Willingdon Hospital, Manali (Kullu district of Himachal Pradesh), Chinchpada Christian Hospital (Nandurbar district of Maharashtra) and Civil Hospital, Sector 45, Chandigarh. All study hospitals were charitable hospitals except in Chandigarh where it was a government facility. The catchment population for these hospitals ranged from 100,000 to 700,000. Site descriptions are summarised in Table 1.

**Table 1:**
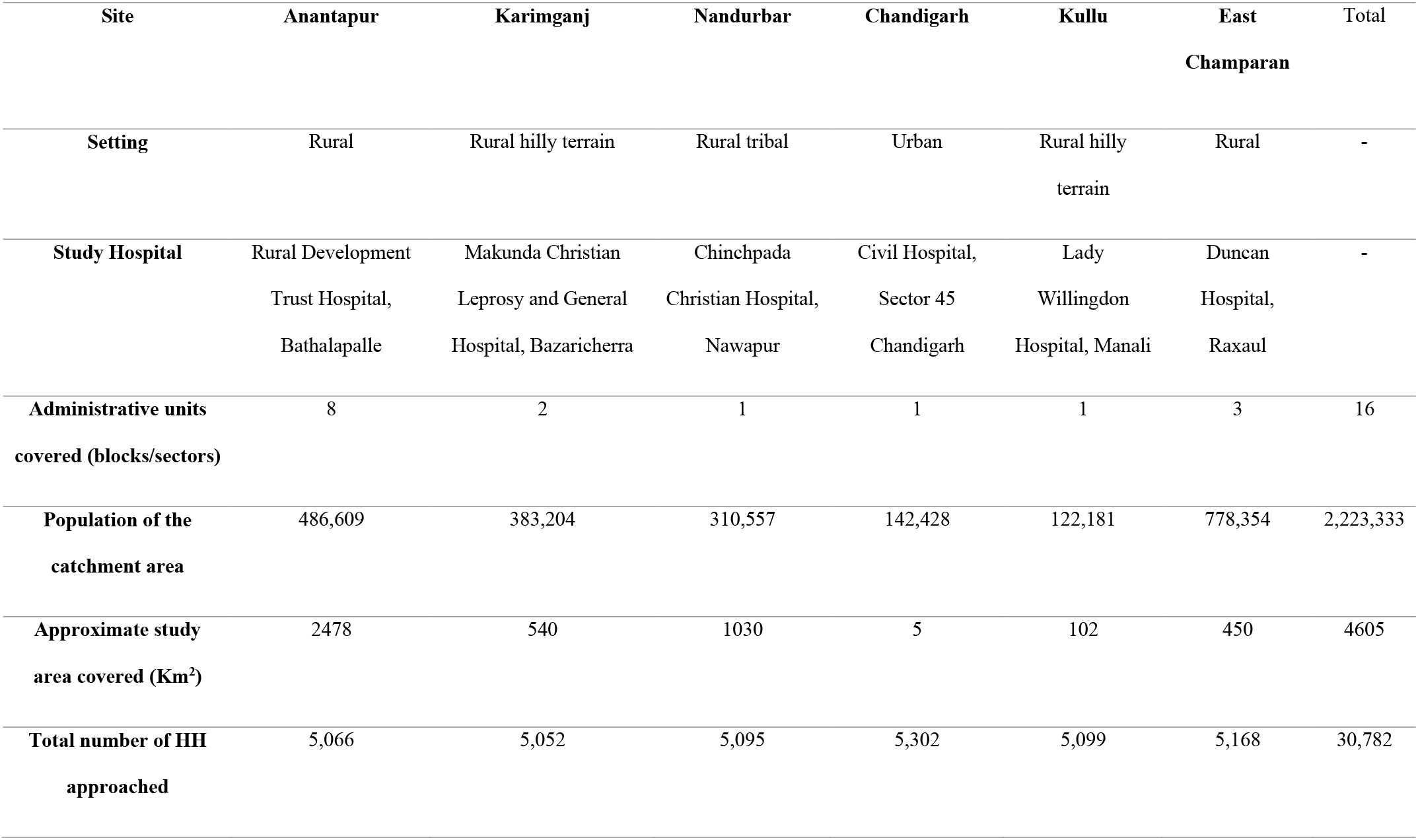
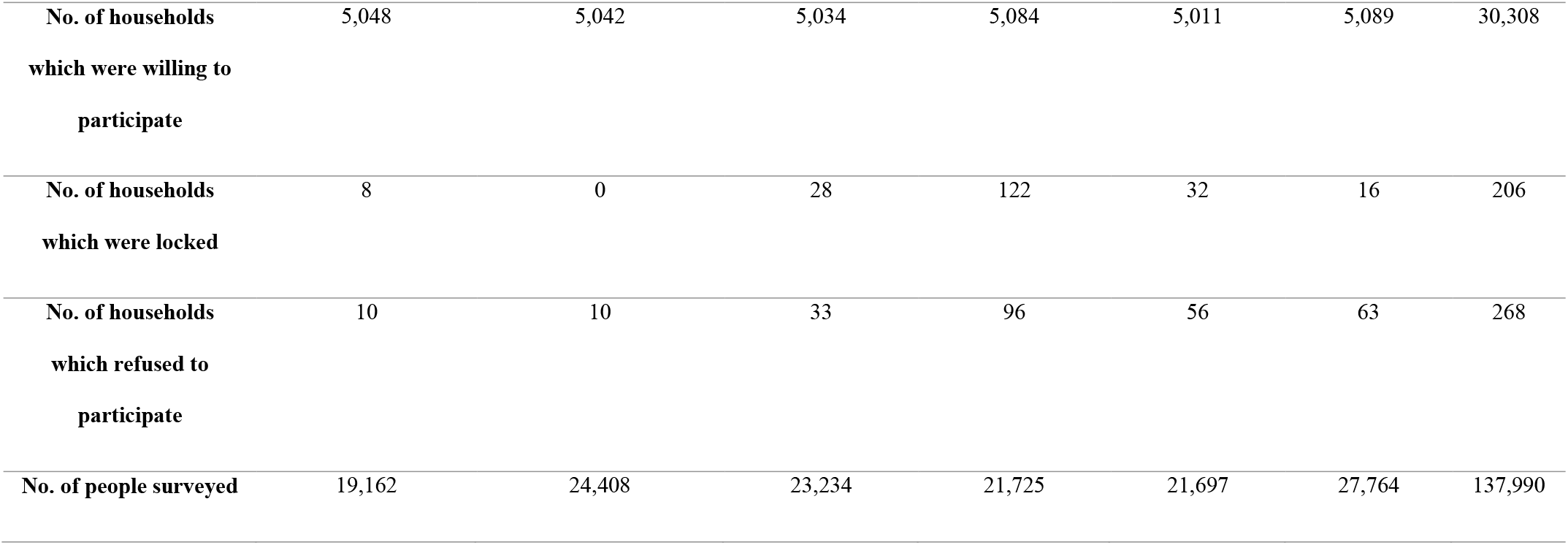
Description of tier 2 surveillance sites and HCUS survey outline.

### Sampling process

The Tier 2 sites were selected, assuming that they were the dominant providers of inpatient medical care in their catchment areas. The catchment area of each of the study hospitals was defined as the geographically contiguous areas from where the most rec**ent 1000 febrile** hospitalisations occurred. This required the review of inpatient hospital records of previous 24 months in each site. Patient addresses were ordered by distance from the study facility, and the Community Development Blocks (CD Blocks) that covered more than 60% of the recent 1000 febrile admissions formed the catchment of the hospital. It was anticipated that about 60% of febrile hospitalisations from the catchment area would occur in the study hospital. The annual incidence of hospitalisation for febrile illness in these settings has been previously estimated at 6/1000 hospitalisation by the National Sample survey Office [19].

We calculated that 150 febrile hospitalisations in the catchment population was required to estimate the proportion hospitalised at the study facility with a ±10% precision and assuming a design effect of 1.5 for intra-familial and village level clustering. Thus we surveyed 25,000 individuals from about 5000 randomly selected households (assuming five persons per household) to identify 150 febrile hospitalisations in each site.

Two rounds of a Healthcare Utilization Survey (HCUS) were conducted in the catchment areas in 2018-2019 with a gap of 6 months between the two to account for any seasonal variations in febrile admissions. In each round, a two-stage sampling process was used. In the first stage, a random sample of 100 primary sampling units (PSU) (wards in urban and villages in rural areas) were selected by probability proportional to size sampling (PPS) technique in each site. Subsequently, systematic random sampling was used to select households within the clusters. From a random start, 50 households were selected from each of the 100 clusters to obtain 5000 household interviews each from six sites.

### Data collection

Data was collected using a questionnaire which was divided into seven sections to collect data on socioeconomic, demographic variables, mortality and morbidity profile, and health-seeking behaviour based on a two week and 12-month recall. Details of any hospitalisations based on a 12-month recall, such as diagnosis, febrile status, treatment facility attended, duration of admission, the severity of illness, and cost of illness, were recorded. Health facility where the patient was admitted was one of the most important questions asked. The survey also collected data on any reported illness in the past two weeks. Details of the diagnosis, antibiotic use, treatment facility, and cost of treatment were collected. The questionnaire also had a section on death, with the diagnosis, febrile status, and the treatment facility used. Data was gathered on a paper-based questionnaire for the first round, and electronically using the “Survey Solutions”, an app developed by The World Bank, for the second round. A responsible member (>15 years of age) of each household was selected as the respondent after obtaining written informed consent. The questionnaire was translated into five regional languages, and then back-translated to check for accuracy. Training modules were developed to train 84 field workers and 12 supervisors (14 field workers and two supervisors per site) who would be responsible for dispensing the questionnaire and primary data collection. The monitoring and evaluation team was also trained separately.

### Monitoring process

Data quality assurance is an organised process intended to ensure the quality of the data collected. This multidimensional process has accuracy, timeliness, accessibility, and comparability as its indispensable components. The survey team was trained to have unambiguous standards for quality checks to adhere to proposed methods to achieve high standards [20,21]. The National Institute of Epidemiology (ICMR-NIE), Chennai was responsible for the monitoring and validation of the survey. The intrinsic hierarchical workflow of the software (Figure 1) allowed quality checks at different points during data collection. Field-workers formed the lowest tier (interviewers), and the data they collected was checked by the supervisors (second tier). The headquarters consisted of the ICMR-NIE team who performed a final check on the data collected for any quality issues. In case any were found at either the supervisor or headquarters tiers, the questionnaires were rejected and the fieldworkers re-contacted the appropriate households to rectify them.

**Figure 1:**
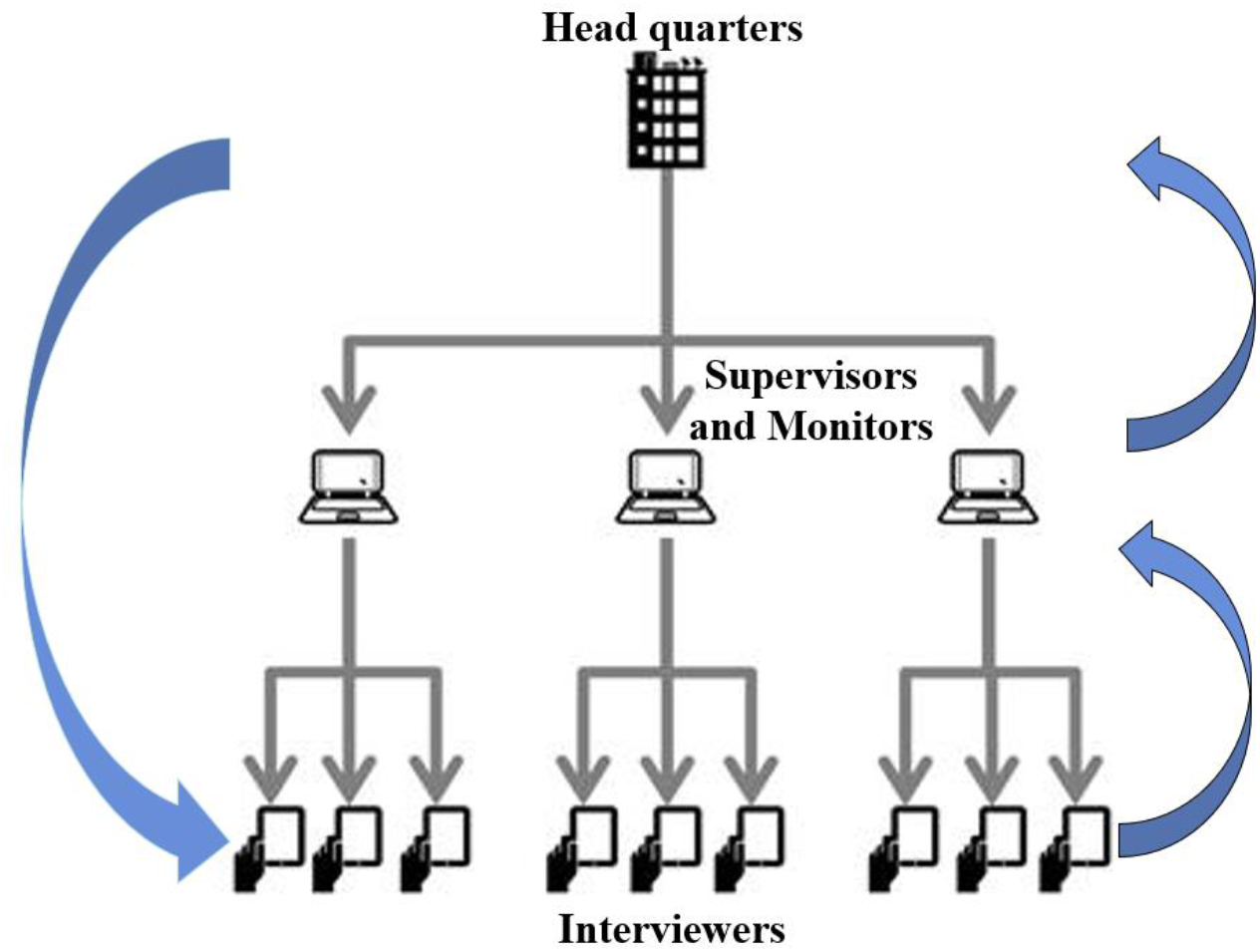
Hierarchical workflow of the healthcare utilisation survey.

Regular field monitoring, along with real-time data monitoring, was executed for each site based on the performance monitoring metrics (Figure 2). The components of the performance monitoring matrix were response rate, time taken for each interview, completeness of data collected, validation by an independent monitor, and comparison with external validator indicators from National Sample Survey (NSS) and Census India 2011. The central monitoring team quantitively measured the performance at each site based on performance scores given to the field workers and clusters separately. Fieldworker scores which could range from 0-1, were based on the proportion of completeness of documentation of selected parameters and on the percentage of adequately timed interviews. Similarly, Cluster scores, which also could range from 0-1, were created based on weights given for the documentation of the number of admissions, deaths, births from the 12-month recall and any reported illness from the 2-week recall, and their comparison with external national validators such as Census of India, 2011 and the NSS 2014 data. Any cluster with a score of less than 0.5 was considered to have an issue with data quality, and feedback and corrective actions were undertaken.

**Figure 2:**
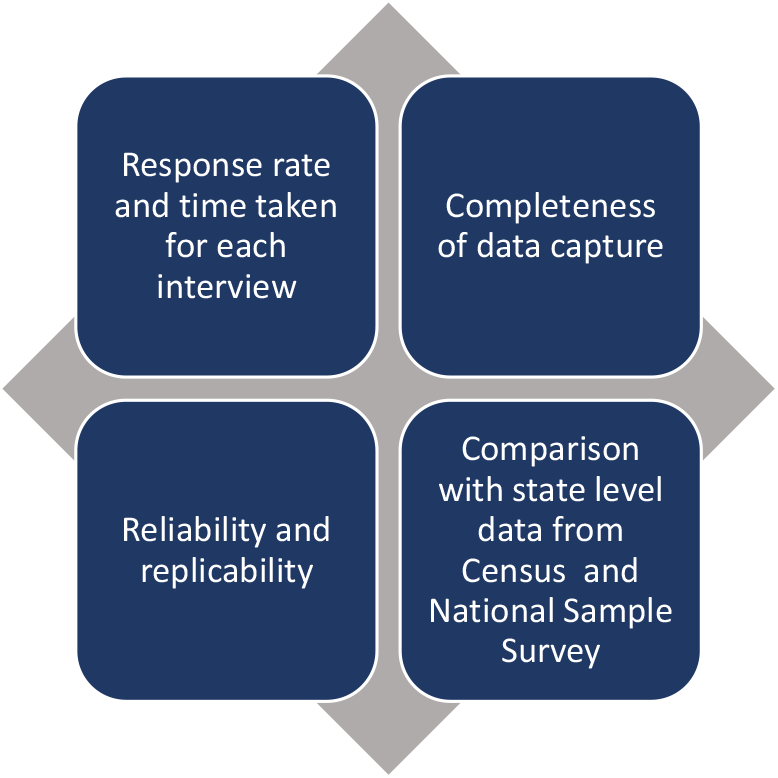
Performance monitoring metrics.

### Statistical analysis

Frequencies and percentages were calculated for the sociodemographic variables. Rates of all-cause and febrile illness and hospitalizations were calculated per 1000 population surveyed. Percentage of the febrile admissions that sought care in the study hospitals were calculated for paediatric age group, adults and overall. Statistical analysis was done using Stata (version 15).

## Results

The results from the second round of HCUS is described here. A total of 30782 households were approached, of which 474 households either refused to participate or were locked. The survey covered a population of 137,990 from the 30,308 households sampled (Table 1). In Anantapur, Nandurbar, and Kullu one-fifth of the surveyed population were children under the age of 15 years, whereas in Karimganj and East Champaran, more than a third of the surveyed population was under 15 years (Table 2). Karimganj and East Champaran had a higher proportion of unmarried people compared to the other sites. All the sites had a higher number of nuclear families than extended/ joint families. Socioeconomic status (SES) was calculated using a modified version of the Kuppuswamy scale [22]. The majority of the households belonged to low socioeconomic strata in all the sites except Kullu where 60% of the households were in the middle SES category. The source of drinking water varied across sites. The primary drinking water source in Anantapur was R/O (Reverse Osmosis) or purified commercial, in Karimganj it was wells, in Nandurbar and East Champaran it was tube wells while in Chandigarh and Kullu it was municipal taps. Open defecation is still prevalent in East Champaran (41%), Nandurbar (26.8%) and Anantapur (13.9%). The survey revealed that access to any health insurance coverage schemes was lowest in Chandigarh (5%) and highest in Anantapur (48%).

**Table 2:**
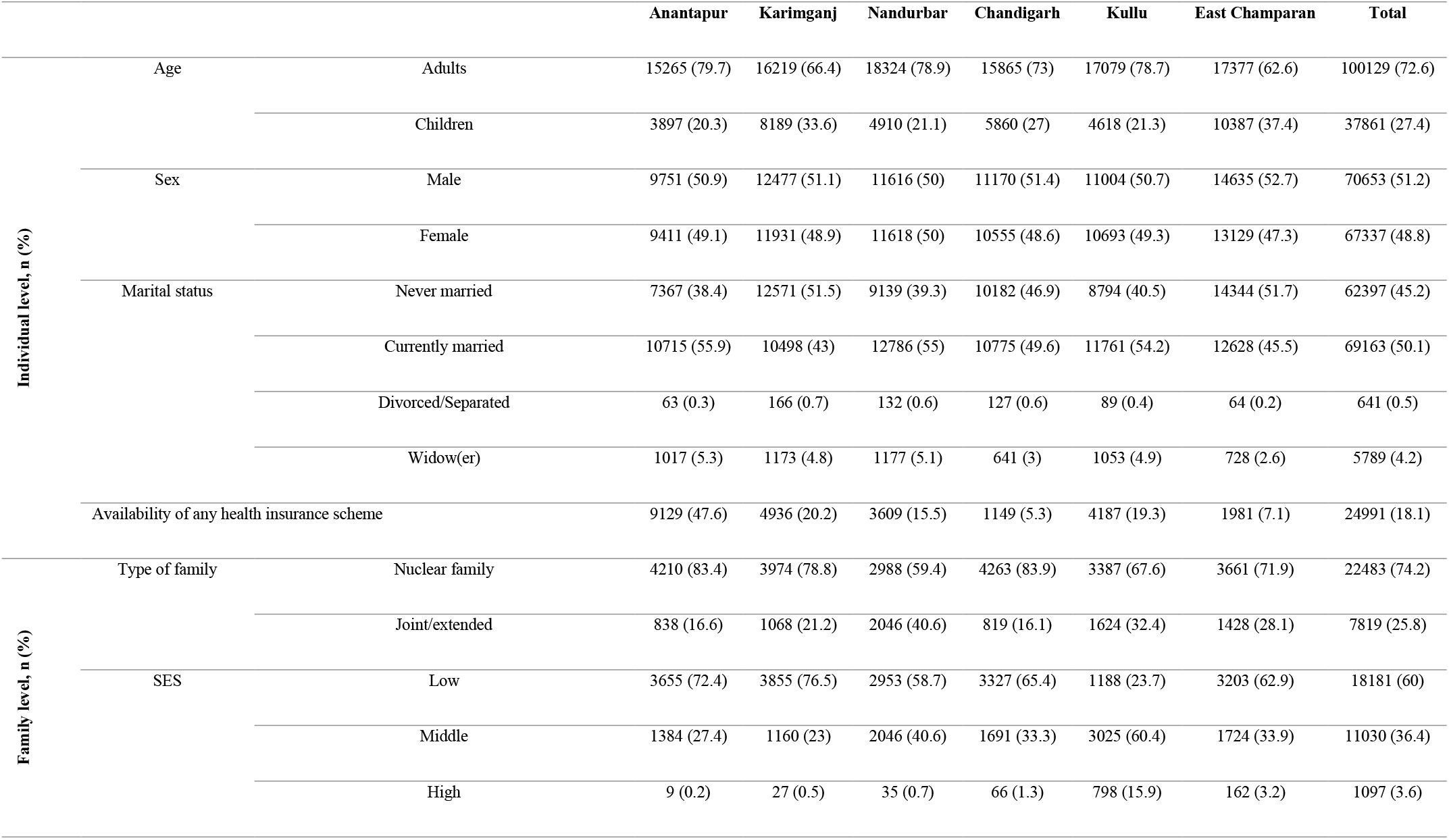
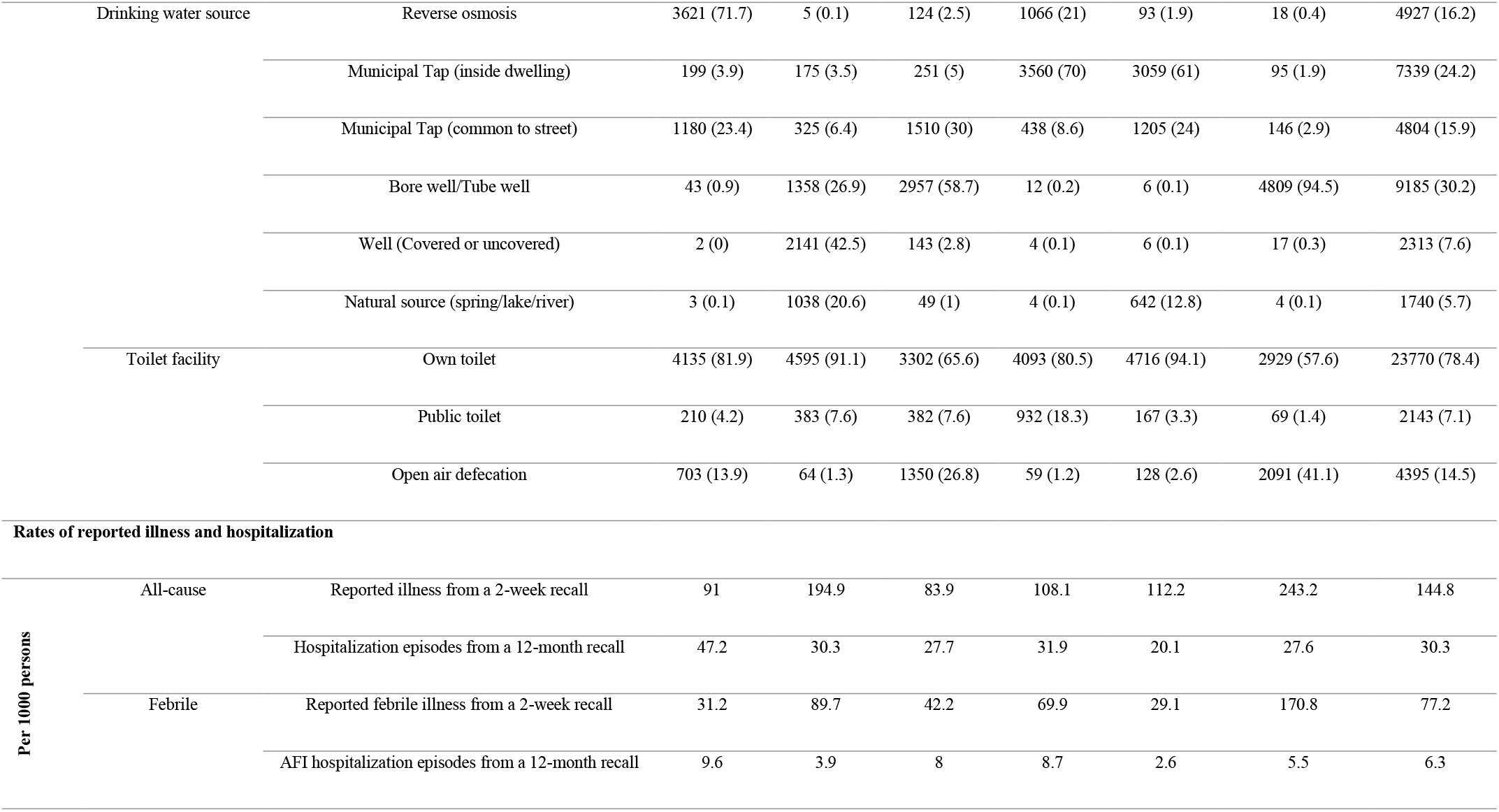
Sociodemographic profile.

Between 84 and 243 individuals per 1000 population reported having any illness requiring either an outpatient visit or was admitted or was managed at home from a 2-week illness recall. Among these, the febrile illness rate from the 2-week recall varied from 29/1000 in Kullu to 170/1000 in East Champaran (Table 2). When we analysed the data on hospitalisations based on a 12-month recall, we found that the hospitalisation rate from any cause ranged from 20/1000 in Kullu to 47/1000 population in Anantapur. Of these, the febrile hospitalisation rate ranged from 2.5 per 1000 in Kullu to 9.6 per 1000 in Anantapur. Approximately a tenth of all the reported deaths were due to a febrile illness.

Of all reported hospitalisations due to any cause 8.5% were among children below 15 years of age. 65.5% of population who were admitted due to any cause were females. The treatment facilities were categorised into private facilities, government facilities, and study hospitals for analysis. Private hospitals including study hospital tended to be used more for admissions for any cause in Anantapur (67.8%) and East Champaran (86.3%), while government facilities were utilised more for admissions due to any cause in Karimganj (50.3%), Nandurbar (53.3%), Kullu (51.5%) and Chandigarh (90%).

When we categorised admissions into febrile causes, non-febrile causes, surgical including trauma causes, and those related to pregnancy and childbirth, we found that in Anantapur and East Champaran, admissions due to non-febrile conditions were highest and was 39.3% and 29.7% respectively. A majority of the hospitalisations were for pregnancy and childbirth in Karimganj (55.4%), Nandurbar (35.5%), Kullu (49.9%) and Chandigarh (38.8%). If pregnancy and childbirth-related admissions were excluded, majority of the admissions were for non-febrile medical conditions in Anantapur (49.5%), Karimganj (40.3%), Kullu (38.4%) and East Champaran (39.2%), and it was for febrile admissions in Nandurbar (45.1%) and Chandigarh (44.3%).

Our results showed that on average, a fifth of all admissions were due to febrile causes. On analysing the febrile hospitalisations alone (Table 3), we found that 77% of the admissions were in adults, and between 23% were among children. The median duration of hospitalisation for these febrile cases across sites was four days. The percentage of febrile admissions among all age groups in the study hospitals was 17.4% in Anantapur, 37.5% in Karimganj, 38.3% in Chandigarh, 9.6% in Nandurbar, 35.7% in Kullu and 23.5 % in East Champaran. When stratified by age groups into adults and pediatric, it was found that the percentage of febrile admissions in the study hospitals was higher for the pediatric age group than the adults in all sites except Chandigarh. Among the pediatric age group, the percentage of febrile admissions in the study hospital was 22.6% in Anantapur, 22.2% in Chandigarh, 11.4% in Nandurbar, 63.6% in Karimganj, 36.4% in Kullu and 34.9% in East Champaran. Proximity to home, low cost of treatment, and good quality care were the main reasons for choosing to approach the study hospitals. We noticed that the number of febrile admissions recorded during the first round of HCUS was more than those recorded in the second round. However, the percentage of febrile hospitalisations in the study hospitals were similar in both the rounds of HCUS across sites.

**Table 3:**
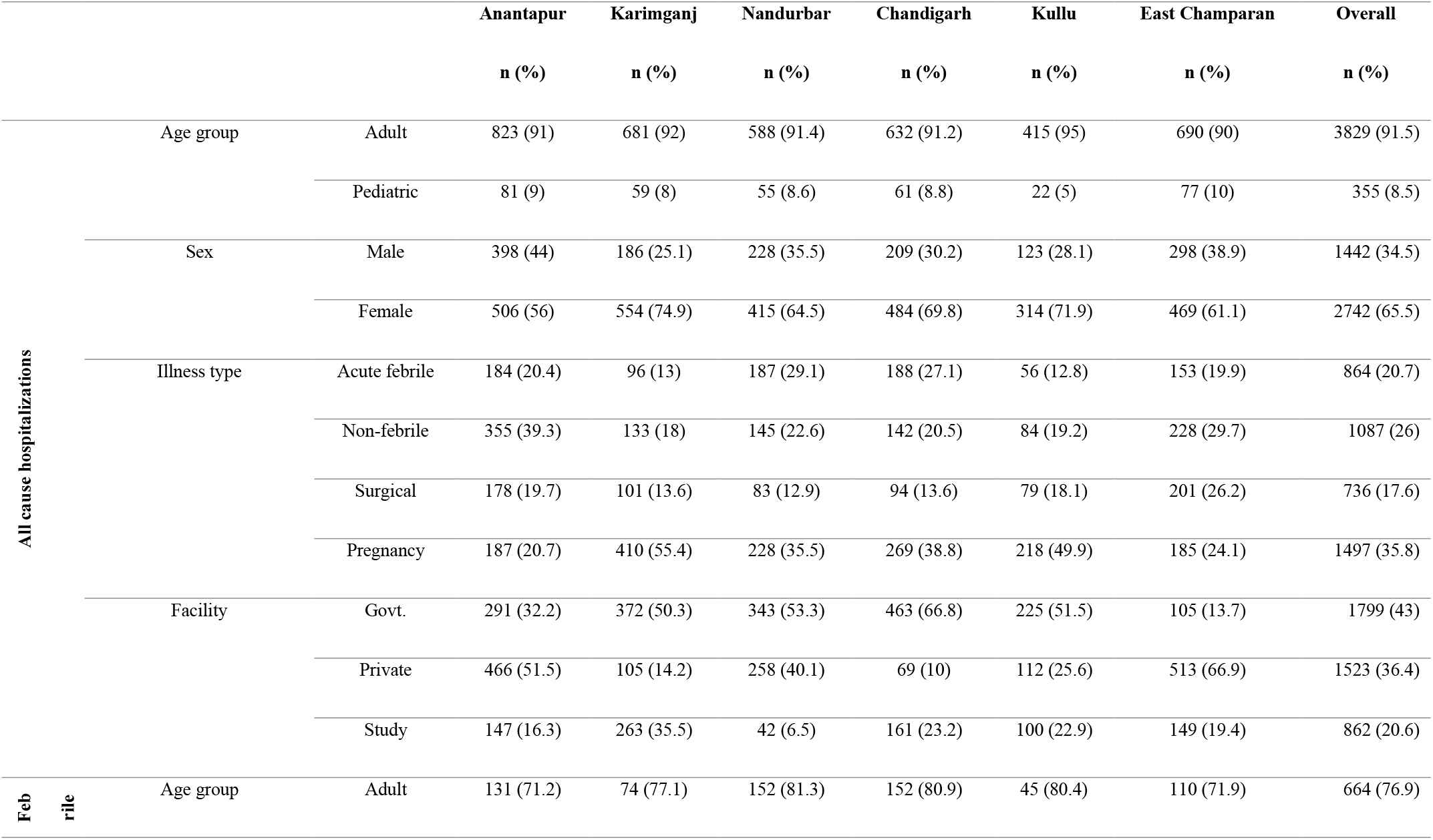
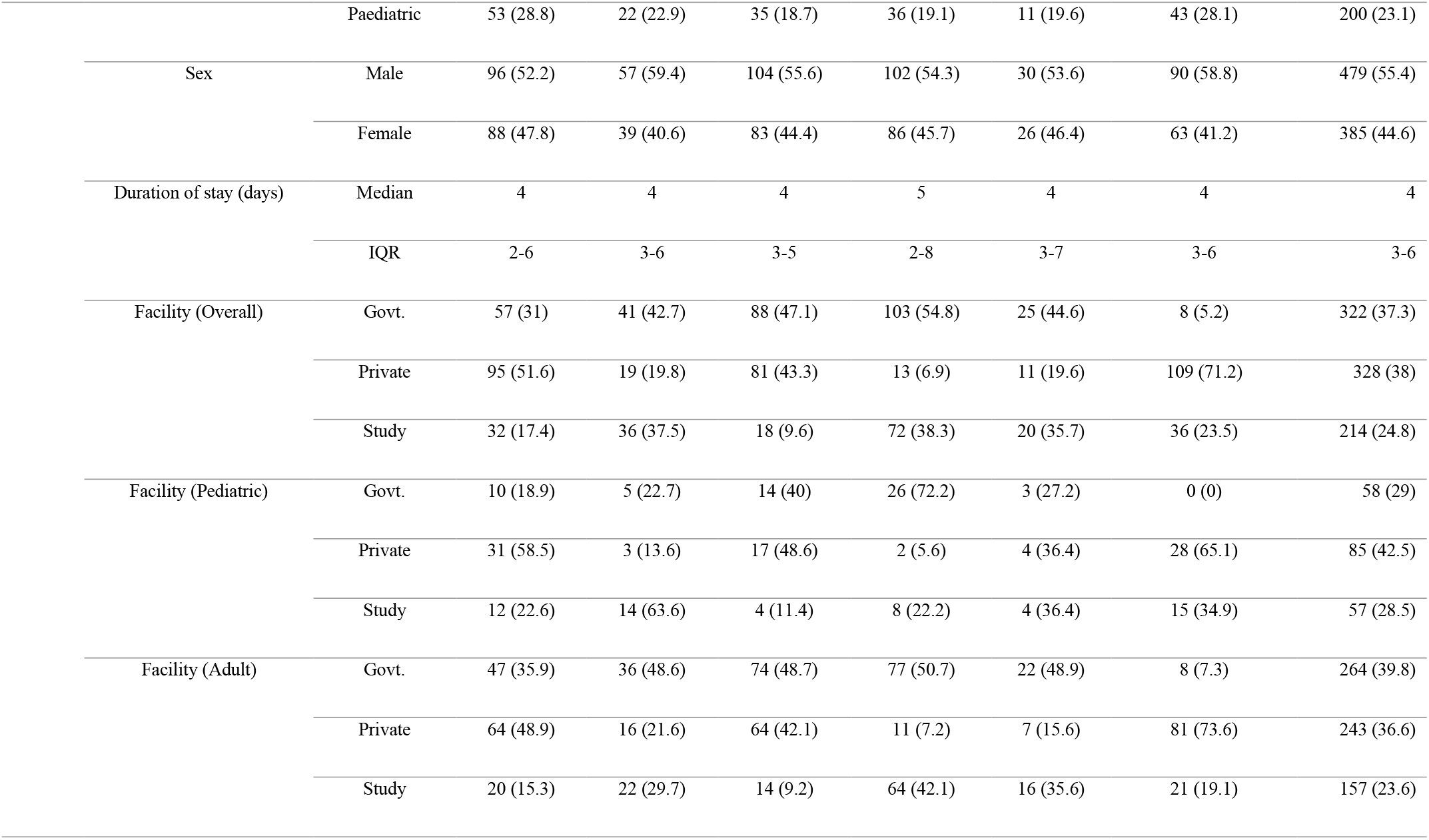
Characteristics of the hospitalized cases from a 12-month recall among the surveyed populations.

### Monitoring

Table 4 summarises the cluster and fieldworker scores at each of the sites. A cluster score of less than 0.5 was considered to have an issue with quality. Of the 600 clusters, 131 clusters had poor quality and had to be resurveyed under supervision. Cluster scores improved after the resurvey. Mean cluster score improved from 0.51 to 0.75 in Anantapur, 0.64 to 0.80 in Karimganj, 0.73 to 0.80 in Chandigarh, 0.67 to 0.72 in Nandurbar, 0.58 to 0.62 in Kullu and 0.71 to 0.75 in East Champaran, after the resurvey. This improvement in scores is statistically significant in all site except Kullu. The mean score for the field workers ranged from 0.64 in Kullu to 0.92 in Nandurbar. The field workers who were not performing well were either retrained or replaced. Figure 3 represents a sample scoring sheet depicting the performance of each cluster in a site.

**Table 4:**
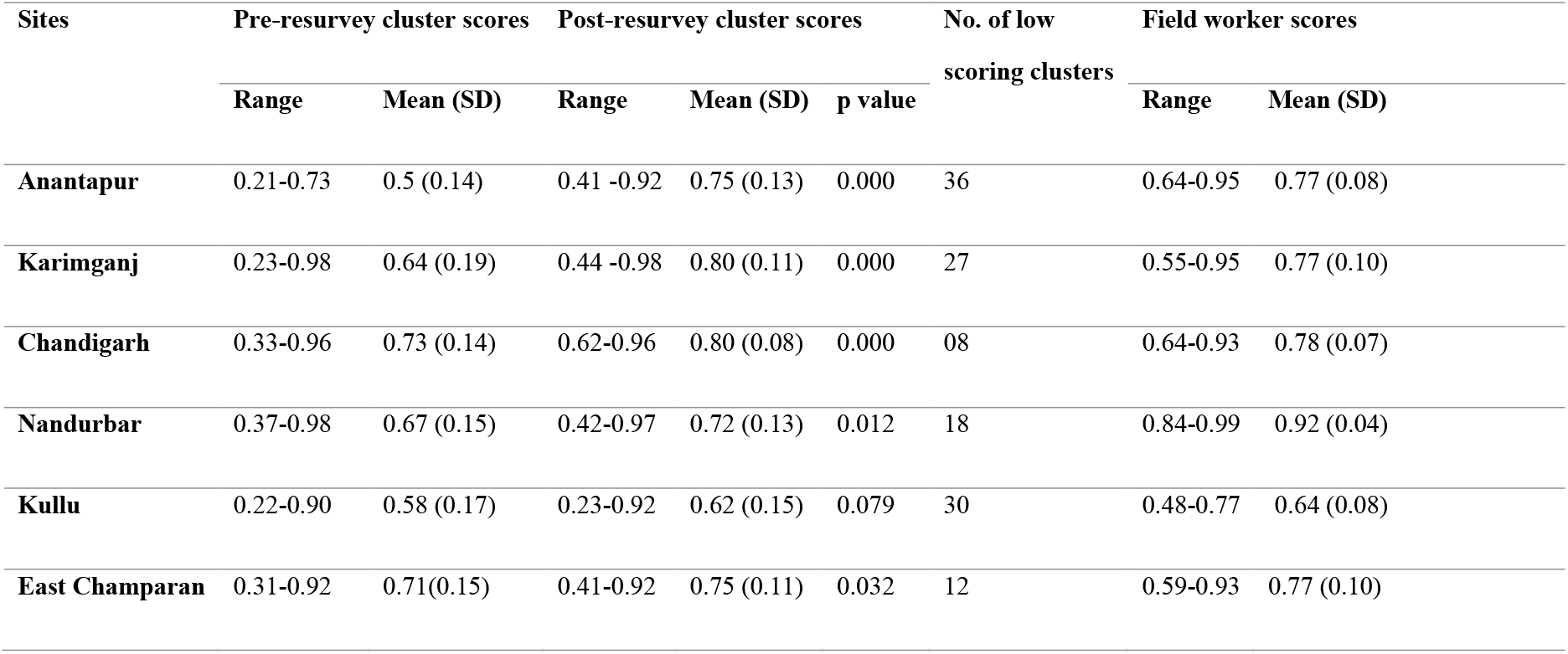
Summary of cluster scores and field worker scores used for monitoring

| Sites | Pre-resurvey cluster scores |  | Post-resurvey cluster scores |  | p value | No. of low scoring clusters | Field worker scores |  |
| --- | --- | --- | --- | --- | --- | --- | --- | --- |
|  | Range | Mean (SD) | Range | Mean (SD) |  |  | Range | Mean (SD) |
| **Anantapur** | 0.21-0.73 | 0.5 (0.14) | 0.41 -0.92 | 0.75 (0.13) | 0.000 | 36 | 0.64-0.95 | 0.77 (0.08) |
| **Karimganj** | 0.23-0.98 | 0.64 (0.19) | 0.44 -0.98 | 0.80 (0.11) | 0.000 | 27 | 0.55-0.95 | 0.77 (0.10) |
| **Chandigarh** | 0.33-0.96 | 0.73 (0.14) | 0.62-0.96 | 0.80 (0.08) | 0.000 | 08 | 0.64-0.93 | 0.78 (0.07) |
| **Nandurbar** | 0.37-0.98 | 0.67 (0.15) | 0.42-0.97 | 0.72 (0.13) | 0.012 | 18 | 0.84-0.99 | 0.92 (0.04) |
| **Kullu** | 0.22-0.90 | 0.58 (0.17) | 0.23-0.92 | 0.62 (0.15) | 0.079 | 30 | 0.48-0.77 | 0.64 (0.08) |
| **East Champaran** | 0.31-0.92 | 0.71(0.15) | 0.41-0.92 | 0.75 (0.11) | 0.032 | 12 | 0.59-0.93 | 0.77 (0.10) |

**Figure 3:**
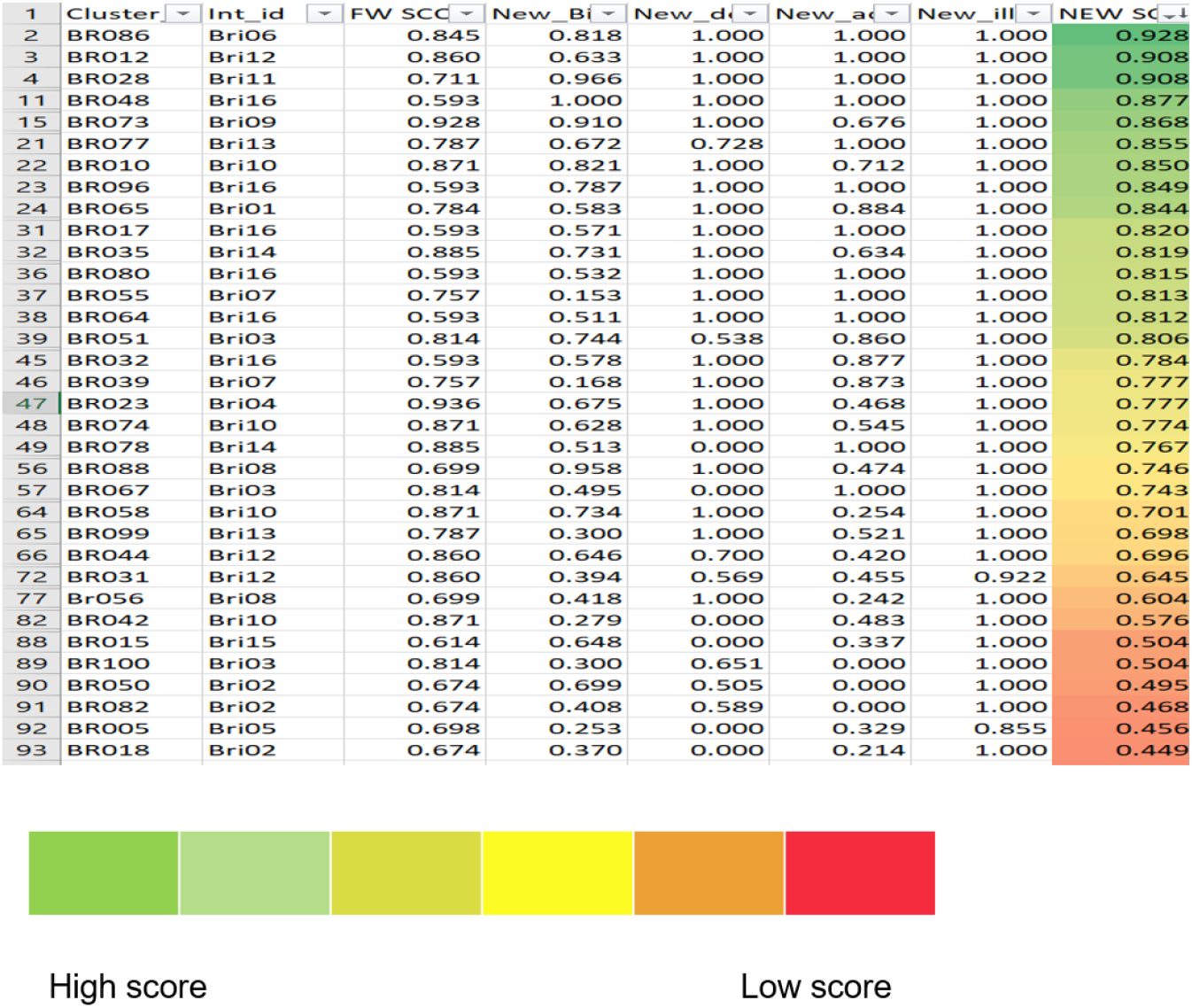
A sample of a scoring sheet depicting the performance of clusters in a site.

## Discussion

The study facility utilisation for febrile hospitalisations in the six sites under this study ranged from 9% to 38%. This observed utilisation rate is lower than the presumed 60%, taken at the time of choosing the facilities for the hybrid surveillance. Low and variable utilisation of the study facility underscores the importance of serially assessing community-based healthcare utilisation to generate adjusted, more accurate estimates of the occurrence of Salmonella infections and other diseases associated with fever [5,11,17,23]. High population density, availability of traditional/different systems of medicine in India, the plurality of private health sector providers which has an almost equal number of qualified doctors and unqualified practitioners, lack of facilities in the public health sectors, etc. could be some of the reasons for low utilisation of any single health facility in a particular site [24]. In areas with multiple healthcare facilities, selecting a single sentinel facility for facility-based surveillance may result in an underestimation of the incidence rates. A solution to this is to select more than one preferred facility from an area to form a network. Such a network of hospitals will improve the sensitivity of the surveillance as more cases will be picked up to contribute to the crude incidence rate. As the percentage of facility coverage increases, the uncertainty around the adjustment factor reduces.

Health is known to have multidimensional facets and have multiple and complex factors that influence it. In a country with widespread differences in socioeconomic status, highly complex health delivery systems, and their distribution, any effort to measure it at a single point will not suffice [25,26]. It is imperative to do serial HCUSs among the catchment population of the sentinel sites due to the seasonality of febrile illness as well as chances of seasonal factors such as harvesting, sowing, rain, and snow affecting access to the hospital. Thus serial HCUSs will avoid incorrect extrapolations and unrepresentative conclusions [5]. The main reasons in our study, for choosing the study hospital for care during an illness that led to admission are proximity to home, low cost, and good quality care. Other studies have suggested that the availability of better infrastructure, quality care, convenient timings, and referral by healthcare providers are the main reasons for preferring private sector hospitals. Availability of qualified healthcare personnel and the availability of free drugs are the main reasons for choosing government facilities for treatment [27]. Other studies in Africa have also reported proximity to the health facility as a significant factor for approaching a specific health facility [23,28].

Though diversity is considered a strength in many aspects, it proved to be a barrier while implementing our extensive surveys with the existing differences in temperature, rainfall, geographical terrain, socio-cultural issues, language, and dialect variations across sites. Finding enough trained field workers who could communicate in the regional language at these sites was a challenge. Access to some of our sites was challenging owing to remote locations and hilly terrain. Kullu posed unique hurdles in the form of floods, washed away roads and bridges, forcing us to halt the survey for a few weeks. Recall bias inherent with a 12 month recall period cannot be ignored. Recall bias could be overcome by selecting a shorter recall period. Refusal to participate, though present, was minimal.

A dedicated monitoring team was constituted to ensure the quality of the data. We had measurable monitoring parameters and transparent monitoring procedures to ensure quality. The tablet-based survey allowed real-time data monitoring which helped in improving the quality of the data gathered. Regular on-site field monitoring was helpful to resolve data quality issues. Developing performance monitoring metrics and identifying key parameters facilitated the development of a scoring system. Monitoring field worker and cluster scores in real-time was beneficial in identifying data quality issues that were subsequently resolved and thus ensured quality.

## Conclusion

Hybrid surveillance is an excellent tool to efficiently and accurately monitor and measure infectious disease burden. When an effective surveillance system is set up, other diseases can also be monitored without much incremental cost. LMICs could utilise this design to collect incidence data which has the potential to guide policy and prioritise the allocation of scarce resources. Annual or bi-annual healthcare utilisation surveys are an integral part of hybrid surveillance to capture any change in the care-seeking behaviour of the catchment area, and true burden estimates are dependent on a well-performed healthcare utilisation survey.

## Supporting information

Conflict of Interest declaration of authors

Strobe checklist

## Data Availability

Data analysis is ongoing and the manuscript is under preparation. Data will be provided upon request.

## Acknowledgements

We would like to express our gratitude to the people who participated in the survey, the field workers and supervisors, the research staff at the National Institute of Epidemiology, Chennai who were involved in the monitoring process, the field staff in the study hospitals of the hybrid surveillance sites, Dr Arun K S and Dr Swathi Krishna of The Wellcome Trust Research Laboratory CMC Vellore for their support during data collection.

## Footnotes

### Meeting where the information was previously presented

Part of the information was presented as a poster titled “Monitoring process of a Healthcare Utilization Survey in six selected sites across India” during ASCODD 2020 held in Bangladesh in January 2020

## References

1. WHO | WHO recommended surveillance standards, Second edition [Internet]. World Health Organization; [cited 2019 Dec 18]. Available from: https://www.who.int/csr/resources/publications/surveillance/WHO_CDS_CSR_ISR_99_2_EN/en/

2. Sharma R, Ratnesh L, Karad AB, Kandpal H, Dhariwal AC, Ichhupujani RL. Communicable disease outbreak detection by using supplementary tools to conventional surveillance methods under Integrated Disease Surveillance Project (IDSP), India. J Commun Dis. 2009; 41(3):149– 159.

3. Oum S, Chandramohan D, Cairncross S. Community-based surveillance: a pilot study from rural Cambodia. Trop Med Int Health. 2005; 10(7):689–697.

4. Technical Contributors to the June 2018 WHO meeting. A definition for community-based surveillance and a way forward: results of the WHO global technical meeting, France, 26 to 28 June 2018 [Internet]. 2019 Jan p. 2019;24(2):pii=1800681. Available from: https://www.ncbi.nlm.nih.gov/pmc/articles/PMC6337056/

5. Bigogo G, Audi A, Aura B, Aol G, Breiman RF, Feikin DR. Health-seeking patterns among participants of population-based morbidity surveillance in rural western Kenya: implications for calculating disease rates. Int J Infect Dis. 2010; 14(11):e967–e973.

6. Sheikh S, Qureshi RN, Raza F, et al. Self-reported maternal morbidity: Results from the community level interventions for pre-eclampsia (CLIP) baseline survey in Sindh, Pakistan. Pregnancy Hypertens. 2019; 17:113–120.

7. O’Donnell O. Access to health care in developing countries: breaking down demand side barriers. Cad Saude Publica. 2007; 23(12):2820–2834.

8. Groce NE, Reeve ME. Traditional healers and global surveillance strategies for emerging diseases. Emerg Infect Dis. 1996; 2(4):351–353.

9. Porter G, Grills N. Medication misuse in India: a major public health issue in India. J Public Health. 2016; 38(2):e150–e157.

10. Bodavala R. Evaluation of Health Management Information System in India. :37.

11. Burton DC, Flannery B, Onyango B, et al. Healthcare-seeking Behaviour for Common Infectious Disease-related Illnesses in Rural Kenya: A Community-based House-to-house Survey. J Health Popul Nutr. BioMed Central; 2011; 29(1):61.

12. Smolinski MS, Crawley AW, Olsen JM, Jayaraman T, Libel M. Participatory Disease Surveillance: Engaging Communities Directly in Reporting, Monitoring, and Responding to Health Threats. JMIR Public Health Surveill [Internet]. 2017 [cited 2019 Dec 17]; 3(4). Available from: https://www.ncbi.nlm.nih.gov/pmc/articles/PMC5658636/

13. Andrews JR, Barkume C, Yu AT, et al. Integrating Facility-Based Surveillance With Healthcare Utilization Surveys to Estimate Enteric Fever Incidence: Methods and Challenges. J Infect Dis. 2018; 218(Suppl 4):S268–S276.

14. Luby SP, Saha S, Andrews JR. Towards sustainable public health surveillance for enteric fever. Vaccine. 2015; 33:C3–C7.

15. Crump JA, Youssef FG, Luby SP, et al. Estimating the Incidence of Typhoid Fever and Other Febrile Illnesses in Developing Countries. Emerg Infect Dis. 2003; 9(5):539–544.

16. D N, Y W, Wc B, et al. Health Care Seeking for Childhood Diarrhea in Developing Countries: Evidence From Seven Sites in Africa and Asia. Am J Trop Med Hyg [Internet]. Am J Trop Med Hyg; 2013 [cited 2020 Mar 14]; 89(1 Suppl). Available from: https://pubmed.ncbi.nlm.nih.gov/23629939/

17. Jordan HT, Prapasiri P, Areerat P, et al. A comparison of population-based pneumonia surveillance and health-seeking behavior in two provinces in rural Thailand. Int J Infect Dis. 2009; 13(3):355–361.

18. Panzner U, Pak GD, Im J, et al. Typhoid fever surveillance in Africa program: Healthcare patterns in febrile study populations. Int J Infect Dis. 2014; 21:246–247.

19. NSSO Government of India. Key Indicators of Social Consumption in India Health. Ministry of Statistics and Programme Implementation; 2014.

20. Üstun TB, Chatterji S, Mechbal A, Murray CJL. Chapter X Quality assurance in surveys: standards, guidelines and procedures. p. 32.

21. Global Adult Tobacco Survey Collaborative Group. Global Adult Tobacco Survey (GATS) Quality Assurance: Guidelines and Documentation Version 2.0. [Internet]. Centers for Disease Control and Prevention; 2010 [cited 2020 Jan 7]. Available from: https://www.who.int/tobacco/surveillance/en_tfi_gats_qualityassurance_v2_final_08dec2010.pdf

22. Deepthi Kattula. Measuring Poverty in Southern India: A Comparison of Socio-Economic Scales Evaluated against Childhood Stunting. PLOS ONE [Internet]. 2016 [cited 2020 Mar 14]; 11(8). Available from: https://journals.plos.org/plosone/article?id=10.1371/journal.pone.0160706

23. Panzner U, Pak GD, Aaby P, et al. Utilisation of Healthcare in the Typhoid Fever Surveillance in Africa Program. Clin Infect Dis Off Publ Infect Dis Soc Am. 2016; 62(Suppl 1):S56–S68.

24. Duggal R. Health care utilisation in India. Health Millions. 1994; 2(1):10–12.

25. Ghosh S, Arokiasamy P. Morbidity in India: Trends, Patterns and Differentials. J Health Stud. 2009;.

26. Dilip TR. Understanding levels of morbidity and hospitalisation in Kerala, India. Bull World Health Organ. 2002; 80(9):746–751.

27. Singh T, Bhatnagar N, Singh G, et al. Healthcare utilisation and expenditure patterns in the rural areas of Punjab, India. J Fam Med Prim Care. 2018; 7(1):39–44.

28. Müller O, Traoré C, Becher H, Kouyaté B. Malaria morbidity, treatment-seeking behaviour, and mortality in a cohort of young children in rural Burkina Faso. Trop Med Int Health TM IH. 2003; 8(4):290–296.

